# Clinical Outcomes of Critically Ill Patients with COVID-19 by Race

**DOI:** 10.1101/2020.09.07.20190074

**Authors:** Fahad Marmarchi, Michael Liu, Srikant Rangaraju, Sara C. Auld, Maria Christina Creel-Bulos, Christine L Kempton, Milad Sharifpour, Manila Gaddh, Roman Sniecinski, Cheryl L. Maier, Fadi Nahab, The Emory COVID-19 Quality and Clinical Research Collaborative (QCRC)

**Affiliations:** Department of Neurology, Emory University School of Medicine, Atlanta, GA; Emory Critical Care Center, Division of Pulmonary, Allergy, Critical Care, and Sleep Medicine, Department of Medicine, Emory University School of Medicine, Department of Epidemiology, Emory University Rollins School of Public Health, Atlanta, GA; Department of Anesthesiology, Division of Critical Care Medicine, Emory University School of Medicine, Atlanta, GA; Department of Hematology and Medical Oncology, Emory University School of Medicine, Atlanta, GA; Department of Anesthesiology, Division of Critical Care Medicine Emory University School of Medicine, Atlanta, GA; Department of Anesthesiology, Emory University School of Medicine, Atlanta, GA; Department of Pathology and Laboratory Medicine, Emory University School of Medicine, Atlanta, GA; Department of Neurology & Pediatrics, Emory University, Atlanta, GA, USA

**Keywords:** COVID-19, mortality, race, African American, outcomes

## Abstract

**Background:** Studies of COVID-19 have shown that African Americans have been affected by the virus at a higher rate compared to other races. This cohort study investigated comorbidities and clinical outcomes by race among COVID-19 patients admitted to the intensive care unit.

**Methods:** This is a case series of critically ill patients admitted with COVID-19 to a tertiary referral teaching hospital in Atlanta, Georgia. The study included all critically ill hospitalized patients between March 6, 2020 and May 5, 2020. Clinical outcomes during hospitalization included mechanical ventilation, renal replacement therapy and mortality stratified by race.

**Results:** Of 288 patients included (mean age, 63 ± 16 years; 45% female), 210 (73%) were African American. African Americans had significantly higher rates of comorbidities compared to other races, including hypertension (80% vs 59%, p=0.001), diabetes (49% vs 34%, p=0.026) and mean BMI (33 kg/m^2^ vs 28 kg/m^2^, p<0.001). Despite African Americans requiring continuous renal replacement therapy during hospitalization at higher rates than other races (27% vs 13%, p=0.011), rates of intubation, intensive care unit length of stay, and overall mortality (30% vs 24%, p=0.307) were similar.

**Conclusion:** This racially diverse series of critically ill COVID-19 patients shows that despite higher rates of comorbidities at hospital admission in African Americans compared with other races, there was no significant difference in mortality.

## Introduction

The coronavirus disease 2019 (COVID-19) has led to death tolls surpassing 700,000 people globally and 160,000 people in the US [1,2]. Studies to date have shown that COVID-19 infects African Americans (AA) at a higher rate compared with other major races in the US; the Centers for Disease Control and Prevention (CDC) has found that AA have a 178.1/100,000 rate of COVID-19, second to only Native Americans and Native Alaskans (221.2/100,000) and significantly higher than that of white Americans (40.1/100,000) [3–6]. Studies have also shown that AA have high frequencies of hypertension, diabetes, chronic kidney disease and obesity, factors associated with poor outcomes from COVID-19 [7–10].

There are limited data assessing racial differences in clinical outcomes of critically ill COVID-19 patients. Cohort studies in Louisiana and Georgia found that race was not associated with hospital mortality after adjusting for differences in sociodemographic and clinical characteristics [5,11]. The objective of our study was to evaluate racial differences and outcomes in a racially diverse cohort of critically ill COVID-19 patients admitted to a major academic hospital.

## Methods

This observational cohort study includes data from hospitalized COVID-19 patients admitted from March 6, 2020 to May 5, 2020 who were admitted to Emory Healthcare, a tertiary referral teaching hospital in Atlanta, Georgia. Patients 18 years or older were eligible for inclusion once positive COVID-19 status was confirmed based on a severe acute respiratory syndrome coronavirus 2 polymerase chain reaction assay. Patient data, including sociodemographic information, clinical data, and laboratory data, were obtained from the electronic medical record (EMR) (Cerner Millenium EMR, Kansas City, MO, USA) and proprietary data collection software developed on the Oracle Apex platform (Redwood City, CA, USA). Codified and continuous data elements were automatically extracted using an internally validated technique from the Millenium EMR. Data elements contained in free-text were manually abstracted by members of the Emory COVID-19 Quality and Clinical Research Collaborative (QCRC) and entered into the Apex platform [12]. For analysis, data from the Apex environment were merged with data directly extracted from the Millenium platform.

Data were collected through June 9, 2020 and analyzed using a chi-square or Wilcoxon rank-sum test for categorical and continuous variables, respectively, with a two-sided p-value of less than 0.05 considered statistically significant (Stata Version 12.1; StataCorp LLC, College Station, TX). This study was approved by the Emory University Institutional Review Board.

## Results

During the study period, 288 patients were admitted to the ICU with a diagnosis of COVID-19. The mean age was 63 (±16) years, 131 (45%) were female, 209 (73%) were AA, 68 (24%) white, 10 (3.4%) were Asian and 1 (0.3%) multiple race. Compared with non-AA, AAs had significantly higher rates of hypertension (80% vs 59%, p = 0.001) and diabetes (49% vs 34%, p = 0.026), and a higher mean BMI (33 vs 28, p<0.001) (Table 1). There were no differences between races in history of coronary artery disease, stroke, chronic kidney disease, chronic obstructive pulmonary disease, asthma, obstructive sleep apnea, HIV or other immunocompromised state.

**Table 1.** Comorbidities in ICU hospitalized patients by race.

| **Admission Characteristics** | **Total n=288** | **AA n=209** | **Other n=79** | **P-Value** |
| --- | --- | --- | --- | --- |
| Age, SD | 63 (16) | 62 (15) | 65 (18) | 0.269 |
| Female, n (%) | 131 (45) | 95 (45) | 36 (46) | 0.986 |
| HTN, n (%) | 214 (74) | 167 (80) | 47 (59) | <0.001 |
| DM, n (%) | 129 (45) | 102 (49) | 27 (34) | 0.026 |
| BMI (n=285), mean (SD) | 32 (8) | 33 (9) | 28 (6) | <0.001 |
| CAD, n (%) | 40 (14) | 25 (12) | 15 (19) | 0.124 |
| Stroke, n (%) | 39 (14) | 26 (12) | 13 (16) | 0.374 |
| CKD, n (%) | 43 (15) | 34 (16) | 9 (11) | 0.300 |
| COPD, n (%) | 19 (7) | 12 (6) | 7 (9) | 0.341 |
| Asthma, n (%) | 28 (10) | 17 (8) | 11 (14) | 0.139 |
| OSA, n (%) | 24 (8) | 18 (9) | 6 (8) | 0.780 |
| HIV, n (%) | 5 (2) | 5 (2) | 0 (0) | 0.328 |
| Other Immunocompromised state, n (%) | 11 (4) | 7 (3) | 4 (5) | 0.501 |

*HTN=hypertension; DM=diabetes mellitus; BMI=body mass index; CAD=coronary artery*
*disease; CKD=chronic kidney disease; COPD=chronic obstructive pulmonary disease;*
*OSA=obstructive sleep apnea; HIV=human immunodeficiency virus*

Mean hospital length of stay was 18 (±12) days and ICU length of stay was 11 (±9) days. Overall, 212 (74%) patients were intubated for a mean of 11 (±8) days; 66 (23%) patients required continuous renal replacement therapy (CRRT) for a mean of 9 (±6) days. There were 82 (29%) patients who died. There was no significant difference in length of stay, intubation rates, length of intubation, and deaths between AAs and non-AA patients (Table 2). CRRT was more frequently required in AA patients (27% vs 13%, p = 0.011) whereas intermittent hemodialysis rates were similar (15% vs 9%, p = 0.153).

**Table 2.**
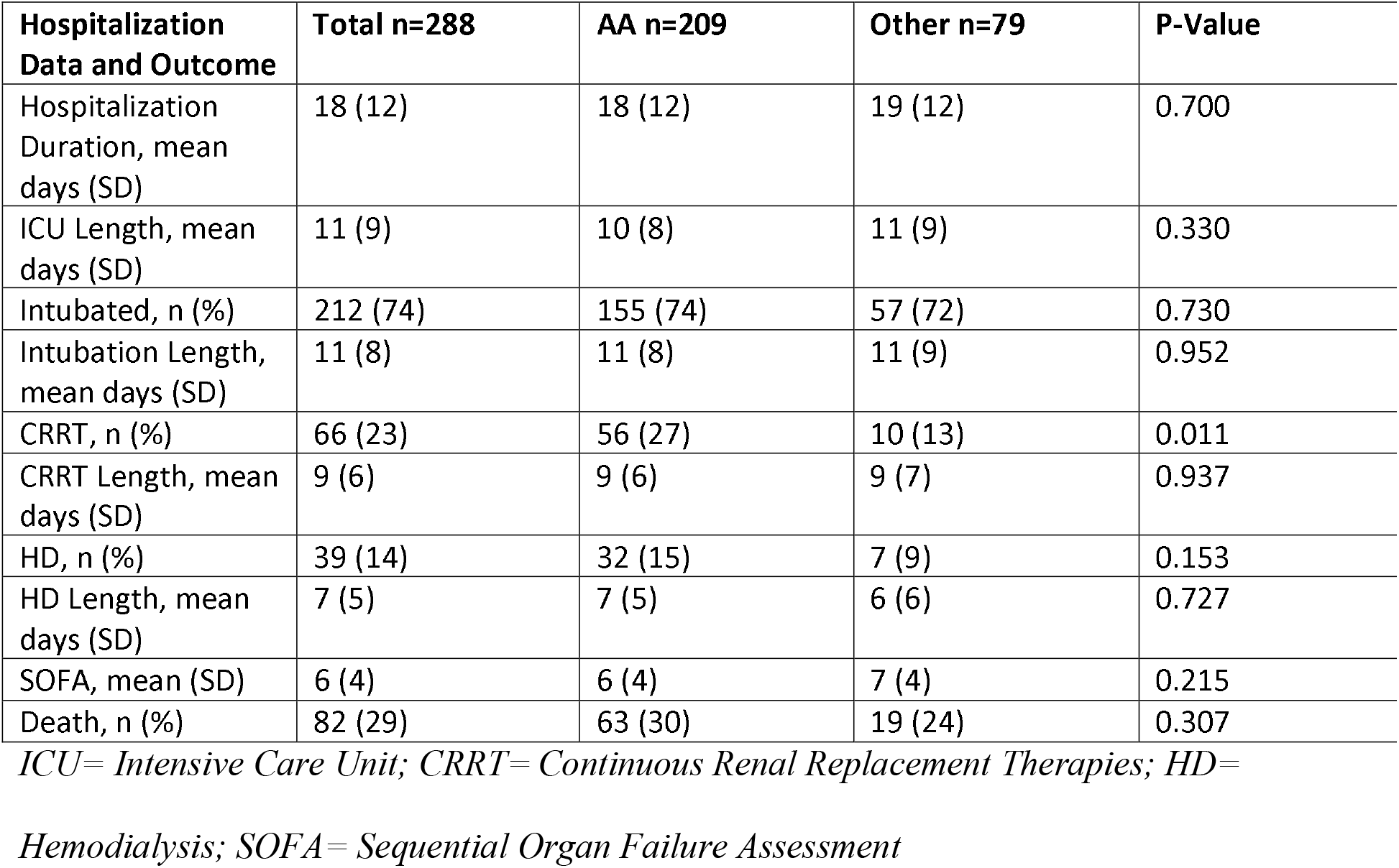
Clinical outcomes in ICU patients by race.

In univariable analysis, factors associated with in-hospital mortality included age, hypertension, coronary artery disease, hospital length of stay (LOS), ICU LOS, intubation rate, CRRT use, hemodialysis treatment duration and BMI but not race. In multivariable analysis, intubation rate, CRRT use, and BMI were associated with in-hospital mortality.

## Discussion

In our racially diverse cohort of critically ill COVID-19 patients including 73% AAs, we found that intubation rate and in-hospital mortality were similar between AA and non-AA patients. This was found despite AAs having higher rates of comorbidities including hypertension, diabetes, and obesity compared to non-AAs. AAs did have higher rates of CRRT and hemodialysis utilization than non-AAs despite having similar rates of chronic kidney disease history on admission. These results are similar to those of a cohort study in Louisiana, which found similar in-hospital mortality rates between AAs and non-AAs despite AAs having higher rates of acute renal failure [5]. In another cohort study from Georgia which included ICU and non-ICU patients, AA patients were found to have similar rates of invasive mechanical ventilation and death compared to non-AA patients [10].

This study has some limitations. Our study was limited to three metropolitan Atlanta hospitals within one academic health system in Georgia and may have limited generalizability to other health care settings. However, our study represents a large cohort of critically ill COVID-19 patients including a large proportion of AA patients. Additionally, this study is limited by being a retrospective analysis of an ICU-specific cohort that does not compare mortality rates outside of an ICU setting. Despite these limitations, our study shows similar in-hospital mortality rates between AAs and other races despite AAs having higher rates of comorbidities at admission and higher rates of RRT in the hospital. Whether these findings are due to pre-hospital social factors [6], differences in the disease manifestation or differences in provider treatments requires further study.

## Conclusion

Despite critically ill AA with COVID-19 presenting with higher rates of comorbidities compared to other races, rates of intubation and mortality were similar at an academic healthcare system in Atlanta, Georgia.

## Article Information

### Corresponding author

Fadi B. Nahab MD Division of Vascular Neurology, Department of Neurology and Pediatrics Emory University School of Medicine, Atlanta, GA

## Data Availability

Clinical data is available upon request for review by informed investigators.

## References

1. Zhang B, Zhou X, Qiu Y, et al. Clinical characteristics of 82 cases of death from COVID-19. PLoS One. 2020;15(7):e0235458.

2. center JHUaMCVR. COVID-19 Dashboard by the Center for Systems Science and Engineering (CSSE) at Johns Hopkins University (JHU). 2020.

3. CenterOfDiseaseControl. Racial & Ethnic Minority Groups 2020; https://www.cdc.gov/coronavirus/2019-ncov/need-extra-precautions/racial-ethnicminorities.html.

4. Millett GA, Jones AT, Benkeser D, et al. Assessing Differential Impacts of COVID-19 on Black Communities. Ann Epidemiol. 2020.

5. Price-Haywood EG, Burton J, Fort D, Seoane L. Hospitalization and Mortality among Black Patients and White Patients with Covid-19. N Engl J Med. 2020;382(26):2534–2543.

6. Cowger TL, Davis BA, Etkins OS, et al. Comparison of Weighted and Unweighted Population Data to Assess Inequities in Coronavirus Disease 2019 Deaths by Race/Ethnicity Reported by the US Centers for Disease Control and Prevention. JAMA Netw Open. 2020;3(7):e2016933.

7. CenterOfDiseaseControl. African American Health 2017.

8. Cooper RS. Genetic Factors in Ethnic Disparities in Health. National Research Council (US) Panel on Race, Ethnicity, and Health in Later Life. 2004;8.

9. Health NIo. Why are some genetic conditions more common in particular ethnic groups?. Genetic Home refrence.

10. Palmer ND, McDonough CW, Hicks PJ, et al. A genome-wide association search for type 2 diabetes genes in African Americans. PLoS One. 2012;7(1):e29202.

11. Gold JAW, Wong KK, Szablewski CM, et al. Characteristics and Clinical Outcomes of Adult Patients Hospitalized with COVID-19 – Georgia, March 2020. MMWR Morb Mortal Wkly Rep. 2020;69(18):545–550.

12. Auld S, Caridi-Scheible M, Blum JM, et al. ICU and ventilator mortality among critically ill adults with COVID-19. Society of Critical Care Medicine. 2020.

